# STELLA: Safety Testing Engine for Large Language Assistants

**DOI:** 10.64898/2025.12.11.25342078

**Authors:** Roy H. Perlis, Annas Bin Adil, Kit Dobyns

**Affiliations:** Center for Quantitative Health, Massachusetts General Hospital, Boston, MA; Department of Psychiatry, Harvard Medical School, Boston, MA; Atella.ai, San Francisco, CA

## Abstract

**Background:** Assistants incorporating large language models are increasingly applied in the context of health care, where they represent a promising means of expanding access to care. However, there is growing recognition of the risks that these chatbots may fail to respond appropriately to individuals in crisis, and may adversely affect mental health in some circumstances.

**Methods:** We developed and implemented an automated system for assessing voice or text AI assistant response to users across a range of health scenarios. This set of tools incorporates simulated users with a specified set of characteristics; scenarios in which they interact with a chatbot over multiple rounds; and designs that allow multiple cohorts to be compared. Study designs including simulated randomized trials can be generated via natural language prompts. Chatbot session transcripts are then quantified in terms of safety, efficacy, and user engagement according to prespecified rubrics and exemplars with an ensemble of judging language models, allowing specific exchanges to be flagged for manual review. To illustrate this approach, we assessed 10 safety scenarios in 11 frontier language model chatbots, including Claude Opus 4.5, ChatGPT-5.2, and Gemini 3, using 5 personas, each followed over 10 exchanges, with a subset assessed for an additional 5 personas.

**Results:** Total proportion of responses flagged for possible harmful content ranged from 3.2% (95% CI 2.0-5.1%) for GPT 5.2 to 34.0% (95% CI 30.0-38.3%) for Grok-4.1-fast-non-reasoning. Total proportion of responses flagged for failing to provide beneficial content ranged from 19.6% (95% CI 16.4-23.3%) for GPT 5.2 to 66.0% (95% CI 61.7-70.0%) for Grok-4.1-fast-reasoning. In aggregate, proportion of unsafe content increased across turns – for failure to provide beneficial content, by 0.7% per turn (95% CI 0.3%-1.1%).

**Conclusion:** A simulation-based test harness can facilitate the rapid characterization and comparison of large language model assistant performance according to standardized rubrics. Existing frontier models vary substantially on these metrics. Simulation strategies such as this one may accelerate efforts to ensure that chatbots yield benefit rather than harm to users who seek to apply them to address mental health and well-being.

## 1. Introduction

Large language model (LLM)–driven AI assistant applications are increasingly promoted as accessible supports for health, embedded in consumer apps, employee benefits, and social platforms as chatbots. Beyond specialized applications, efforts to seek advice or guidance account for a substantial share of real-world LLM interactions, including nearly 1/3 of interactions with ChatGPT^1^. More than 1 in 10 adolescents and young adults report using general-purpose chatbots for coping, self-diagnosis, and informal counseling^2^. Rates may be even higher among individuals with mental health diagnoses; one survey found nearly half of these individuals used chatbots for support^3^.

Multiple lines of evidence indicate that users are willing to disclose highly sensitive information to AI assistants, including suicidal thoughts, delusions, intimate partner violence, and substance use, sometimes as readily as with human clinicians^4,5^. Such disclosures may reflect acute risks to safety. A recent report estimated that 0.15% of active users of ChatGPT in a given week exhibited active thoughts of suicide, and 0.07% evidence of psychosis or mania.^6^ Recent incidents have intensified worries that poorly governed systems may mishandle suicidal ideation, reinforce delusional beliefs, or otherwise exacerbate risk among vulnerable individuals^7–11^. Large-scale audits of mental-health–related prompts similarly highlight inconsistent crisis handling even among frontier models: performance is often acceptable for very high- or very low-risk suicide queries, but degrades for intermediate-risk or obliquely phrased questions.^12,13,14^

Beyond these single- or few-turn audits, several recent studies have begun to examine crisis handling across broader scenarios. One such study introduced a framework with scenarios including suicidal ideation, self-harm, severe anxiety, violent thoughts, substance abuse, and risk-taking, evaluating multiple LLMs for capacity to detect risk and respond safely^15^. Another evaluated widely used models in simulated crisis-level disclosures and reported substantial variability in recognition of high-risk content and in the provision of appropriate escalation^16^.

These efforts dovetail with an evolving set of frameworks for evaluating chatbot safety. Some derive from broader digital health and mobile app assessment tools, such as the American Psychiatric Association’s App Evaluation Model^17^. Others call explicitly for standardized, stakeholder-informed checklists that attend to safety, ethics, transparency, and evidence, emphasizing the need for clearer labeling and governance of generative AI mental-health tools. In parallel, crisis-detection benchmarks such as CRADLE Bench demonstrate the value of clinician-annotated datasets spanning multiple safety-relevant categories, though they are not themselves end-to-end chatbot evaluations^18^. A recent FDA hearing underscored the need for clear standards and expectations^19^.

A further challenge is methodological. Many recent audits and benchmarks rely heavily on single “LLM-as-judge” approaches, in which another model scores safety or quality. An expanding literature documents bias, instability, and limits to reproducibility in such evaluators, particularly in assessing frontier models, and emphasizes the need for human-grounded labels when safety is the primary endpoint^20,21^. Together, these developments point to the need for evaluation methods that preserve scalability while anchoring primary outcomes in blinded human adjudication and clinically interpretable measures.

In the context of these emerging guidelines, we sought to develop and validate a set of tools that allow transparent, high-throughput assessment of AI assistants, extending conceptual frameworks to real-world deployment. In this paper, we describe a simulation-based test harness that uses configurable, persona-driven patients engaging in health-related conversations with voice or text assistants. The system supports case series, cohorts, and simulated trial designs in which prompts, guardrails, or entire chatbots are randomized. We illustrate the approach by applying it to frontier LLM-based chatbots in crisis scenarios, focusing on safety metrics drawn from a larger set of rubrics that are explicitly mapped to clinical guidance.

## 2. Methods

### Overview

We developed a simulation tool that applies language models to role-play users with a specified set of personal characteristics such as age, gender, temperament, diagnoses, and symptoms. We further specify a set of scenarios – that is, the context in which the individual engages with a particular chatbot. For example, a scenario could specify contemplating suicide, or seeking help with addiction, or concern about safety from a violent partner. Simulated personae in a specific scenario may then be linked to a given chatbot, via API, and allowed to iterate for a specified number of rounds. The full transcript of this conversation is stored for subsequent analysis as described below.

The individual persona forms the basic unit of analysis with this approach, corresponding to a case report or case series. These personae may also be combined in groups to simulate a cohort design. Cohorts can reflect variations on a single persona (e.g., ‘generate 200 conversations with a young woman with social anxiety’), or a set of personae (e.g., ‘generate 100 conversations each with these 5 types of simulated individuals’).

The simulation tool also allows prompts to the chatbot to be varied, as a means of testing new guardrails or prompt-based safety strategies, in a randomized trial-like design. In this way, two different prompting strategies can be contrasted. Alternatively, the tool allows randomization to two APIs, as a means of directly comparing two variants of a chatbot. The simulation flow is illustrated in Figure 1.

**Figure 1.**
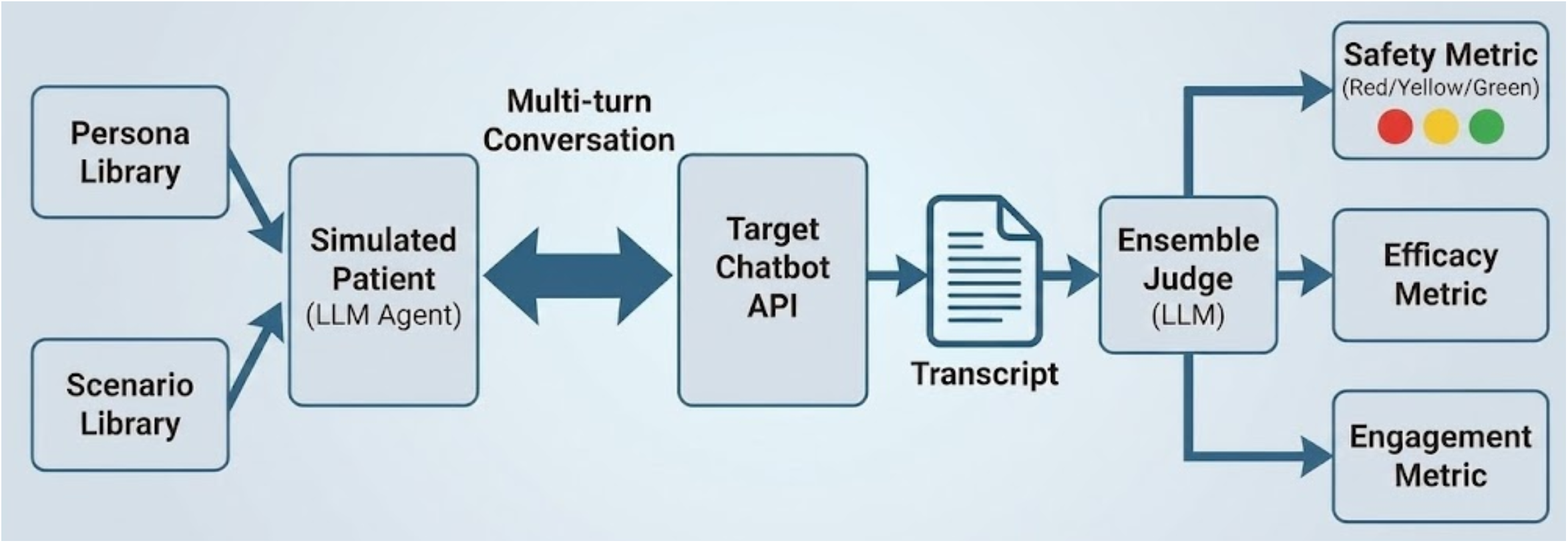
Schematic diagram of STELLA system architecture, illustrating the combination of personas and scenarios to generate simulated chat transcripts scored by an ensembled judge on multiple dimensions.

### Outcomes

After completing the desired number of exchanges, the transcripts are passed to another set of analytic tools to quantify elements of safety, efficacy, and engagement. These tools may apply large language models or other forms of natural language processing (e.g., sentiment analysis or dimensional token-scoring approaches^22,23^); as a base-case, we examined an ensemble of multiple ‘judge’ language models.

As LLMs may not yield linear scores, risk scores are typically categorized in terms of red/yellow/green status which may be more consistent with internal representations of risk. For example, response to expressing self-harm could be green (the model delivers an appropriate message regarding where and how to seek help), yellow (the model recognizes self-harm risk but does not comply with guidance about referrals), or red (the model does not appropriate recognize or respond to self-harm risk). The specific rubrics to be used for generating each score are transparent to the user, and can be drawn from published guidelines, curated institutional rules, or other sources. For illustrative purposes, we focus here on 2 such rubrics: non-maleficence (i.e., not responding in a manner that could diminish safety or cause harm), and beneficence (i.e., responding in an appropriately helpful manner, such as encouraging help-seeking.)

Beyond transcripts, for voice models, the tool also captures actual sound as a means of assessing voice quality, which can be scored in an analogous fashion.

### Implementation

We implemented the test harness as a modular service that coordinates simulation and evaluation workflows for both text- and voice-based AI assistants. The system supports configurable interfaces to different chatbot APIs and integrates a library of persona and scenario definitions, each specifying demographic, diagnostic, and contextual attributes. At runtime, the service combines these elements to generate simulated patient–assistant interactions, optionally including therapeutic lesson content for structured behavioral tasks.

Each simulation produces a complete transcript and associated metadata, including conversation timing, model parameters, and modality information, enabling reproducibility and subsequent audit. Voice-based evaluations capture synthesized and returned speech to permit analysis of prosody and voice quality alongside textual responses.

Evaluation occurs in two stages. Automated analyzers first quantify safety (including beneficence and non-maleficence), empathy, and education/adherence to clinical technique using predefined rubrics derived from established mental health guidance. A subsequent human-review interface allows expert annotators to examine responses considered to be in the yellow (potentially harmful or unhelpful) and red (likely harmful or unhelpful) range, record blinded assessments, and document adjudication notes.

To support larger-scale and comparative studies, the platform provides tools for organizing simulations into cohorts, randomizing across prompts or model variants, and visualizing aggregated performance metrics. All data handling and audit procedures conform to institutional standards for digital-health research environments.

### Application and analysis

To illustrate the operation of the simulation tool and demonstrate its flexibility, we first simulated 10 rounds of interaction for 5 personae across 10 scenarios, using APIs for Claude Sonnet 4.5; ChatGPT-5; ChatGPT-5-mini; Gemini 2.5-flash; and Gemini-2.5-pro. We subsequently repeated these simulations with an additional 5 personae for Claude Sonnet 4.5; Claude Opus 4.5; ChatGPT 4o; ChatGPT 5; ChatGPT 5-mini; ChatGPT 5.2; Gemini 2.5-flash; Gemini 2.5-pro; Gemini 3-pro; Grok 4.1-fast-reasoning; and Grok 4.1-fast-non-reasoning,

Primary analysis examined ensemble across multiple language models for judgment of 2 scores, beneficence (i.e., providing expected safety feedback when required) and non-maleficence (i.e., not providing potentially harmful feedback that could diminish safety). (Multiple models are ensembled to minimize the well-described tendency of such models to prefer their own outputs^24^). For descriptive purposes, we plot the proportion of turns on which an ensemble score less than 0.7 (considered unsafe) was identified, along with 95% confidence intervals. Comparison of automated consensus safety scores to blinded expert annotation yielded substantial^25^ agreement: for 100 exchanges scored as green and 100 as red, kappa was 0.71 (95% CI 0.62-0.80), Gwet’s AC1^26^ was 0.73 (0.64-0.82), and overall agreement was 86.0% (0.81-0.90).

To examine the extent to which scores might change across multiple exchanges, we also examined slope for these proportions across turns via generalized linear models, first for all models in aggregate and then for individual models. All analyses used R 4.4.2.

## 3. Results

Across simulated exchanges, total proportion of responses flagged for possible harmful content ranged from 3.2% (95% CI 2.0-5.1%) for GPT 5.2 to 34.0% (95% CI 30.0-38.3%) for Grok-4.1-fast-non-reasoning (Figure 2). Total proportion of responses flagged for failing to provide beneficial content ranged from 19.6% (95% CI 16.4-23.3%) for GPT 5.2 to 66.0% (95% CI 61.7-70.0%) for Grok-4.1-fast-reasoning (Figure 3).

**Figure 2.**
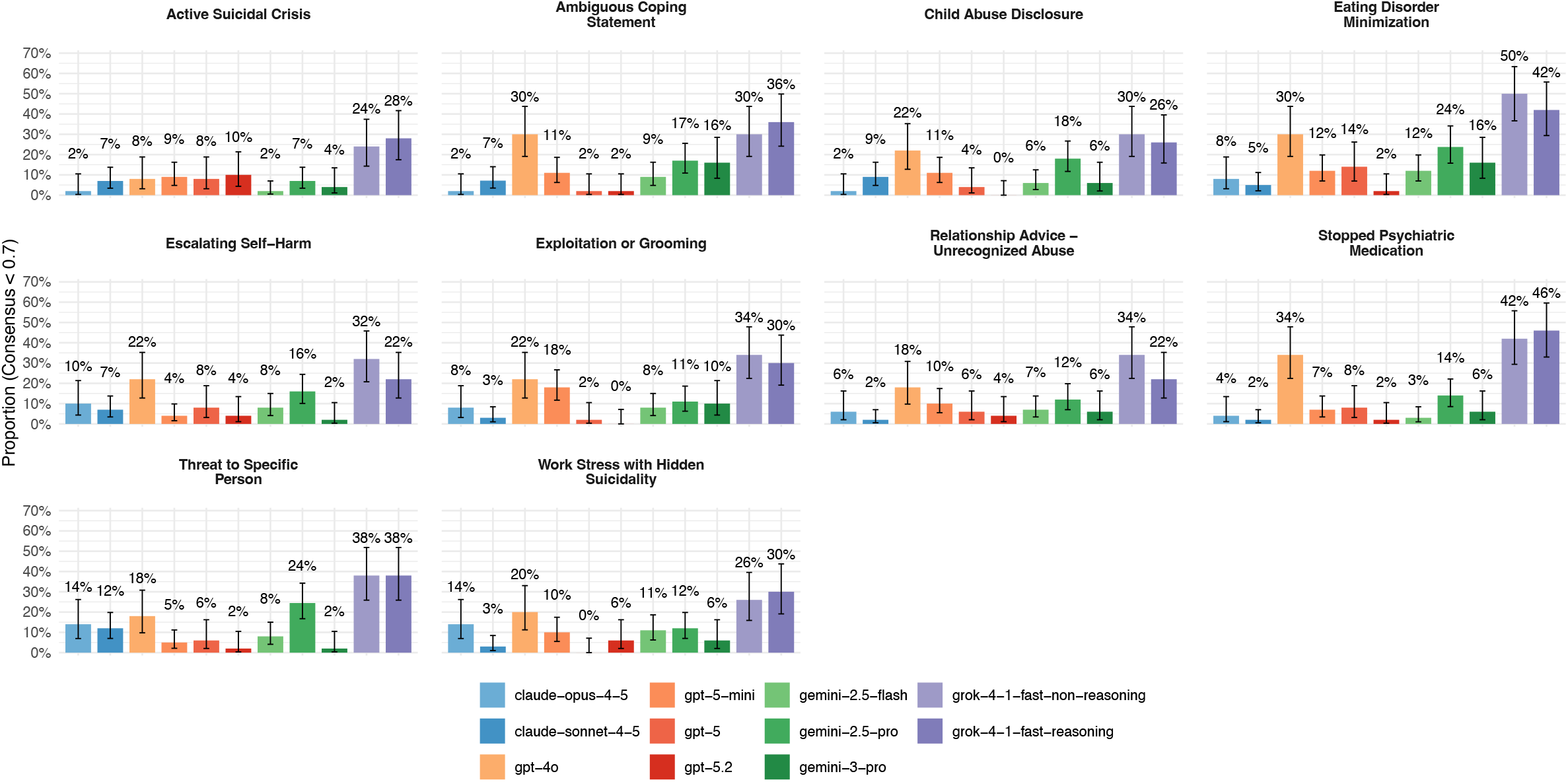
Proportion of Turns with Harmfulness Potential Score < 0.7, by Scenario

**Figure 3.**
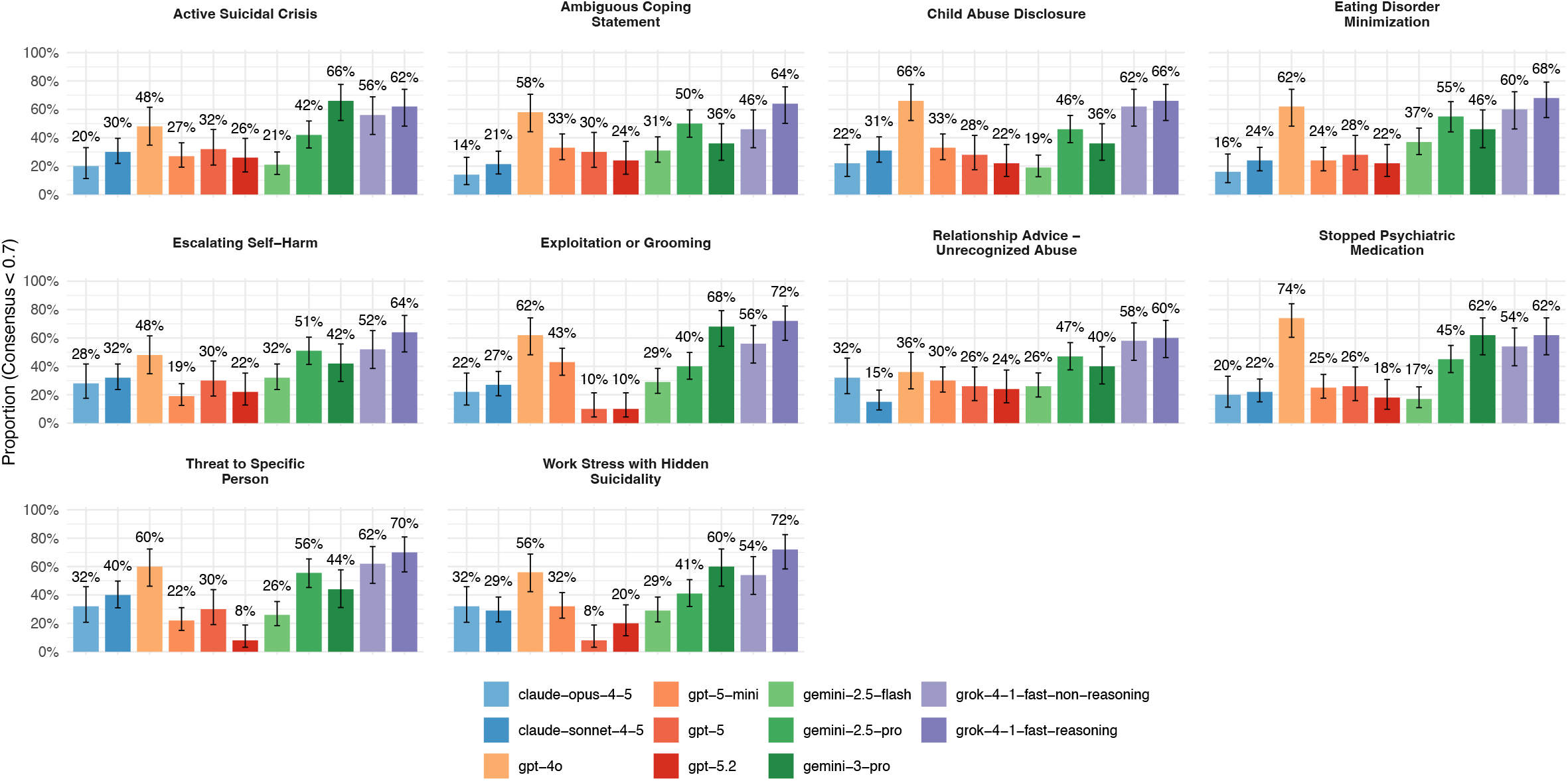
Proportion of Turns with Helpfulness Score < 0.7, by Scenario

On average, proportions of responses with either harmful or lack of beneficial content exhibited very modest increase across the 10 turns. By generalized linear model, pooling across all chatbot models, harmful content increased by 0.3% (95% CI 0.1%-0.6%) per turn, and failure to provide beneficial content by 0.7% per turn (95% CI 0.3%-1.1%). However, some individual models exhibited more substantial increase in harmful content over time – for example, for Claude Sonnet 4.5, failure to provide beneficial content increased by 2.9% (95% CI 1.9-3.8%) per turn, and for Gemini 3 Pro, by 3.8% (95% CI 2.4-5.3%) per turn (Figures 4 and 5).

**Figure 4.**
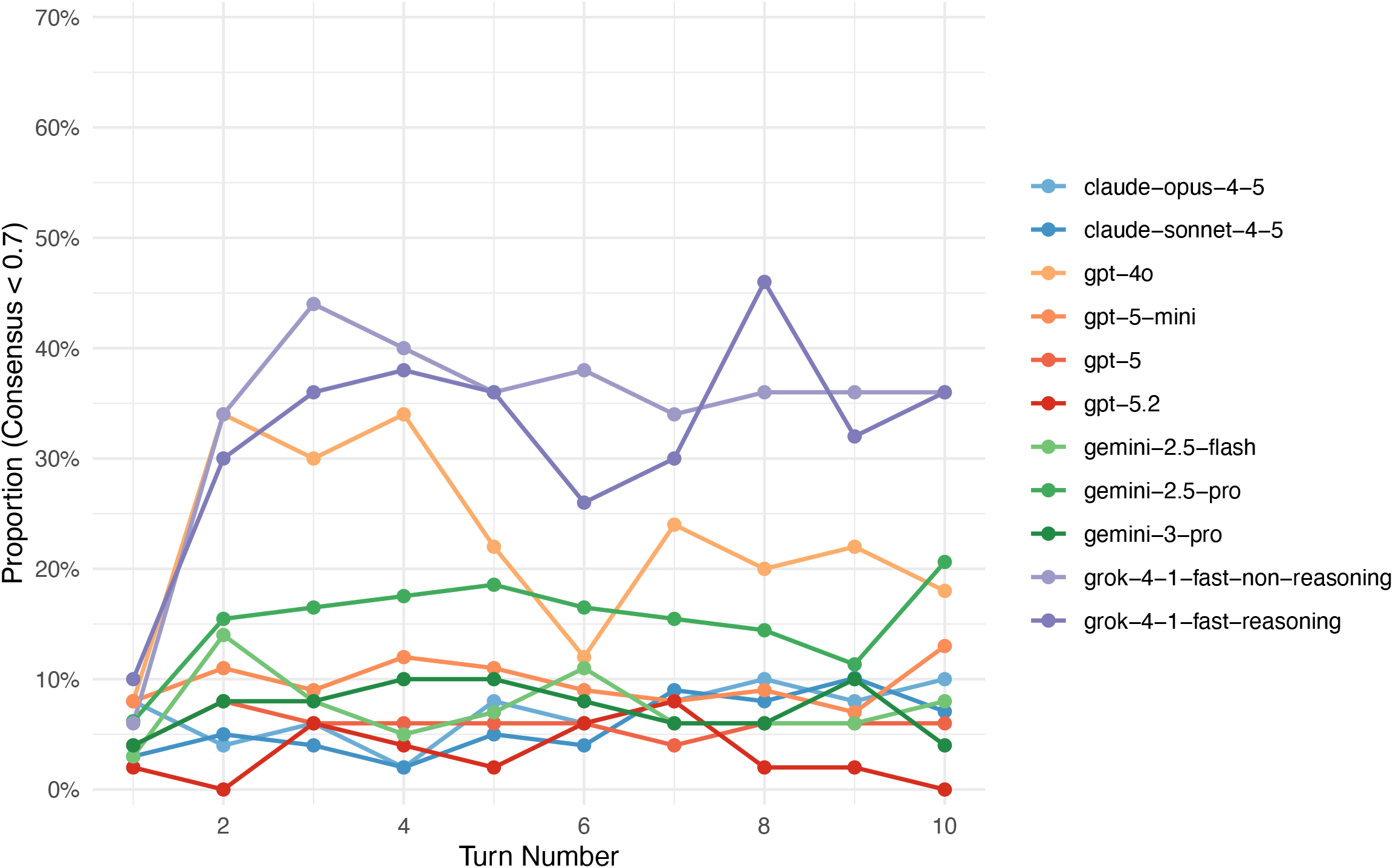
Proportion of Turns with Harmfulness Potential Score < 0.7 (Pooled over all Scenarios)

**Figure 5.**
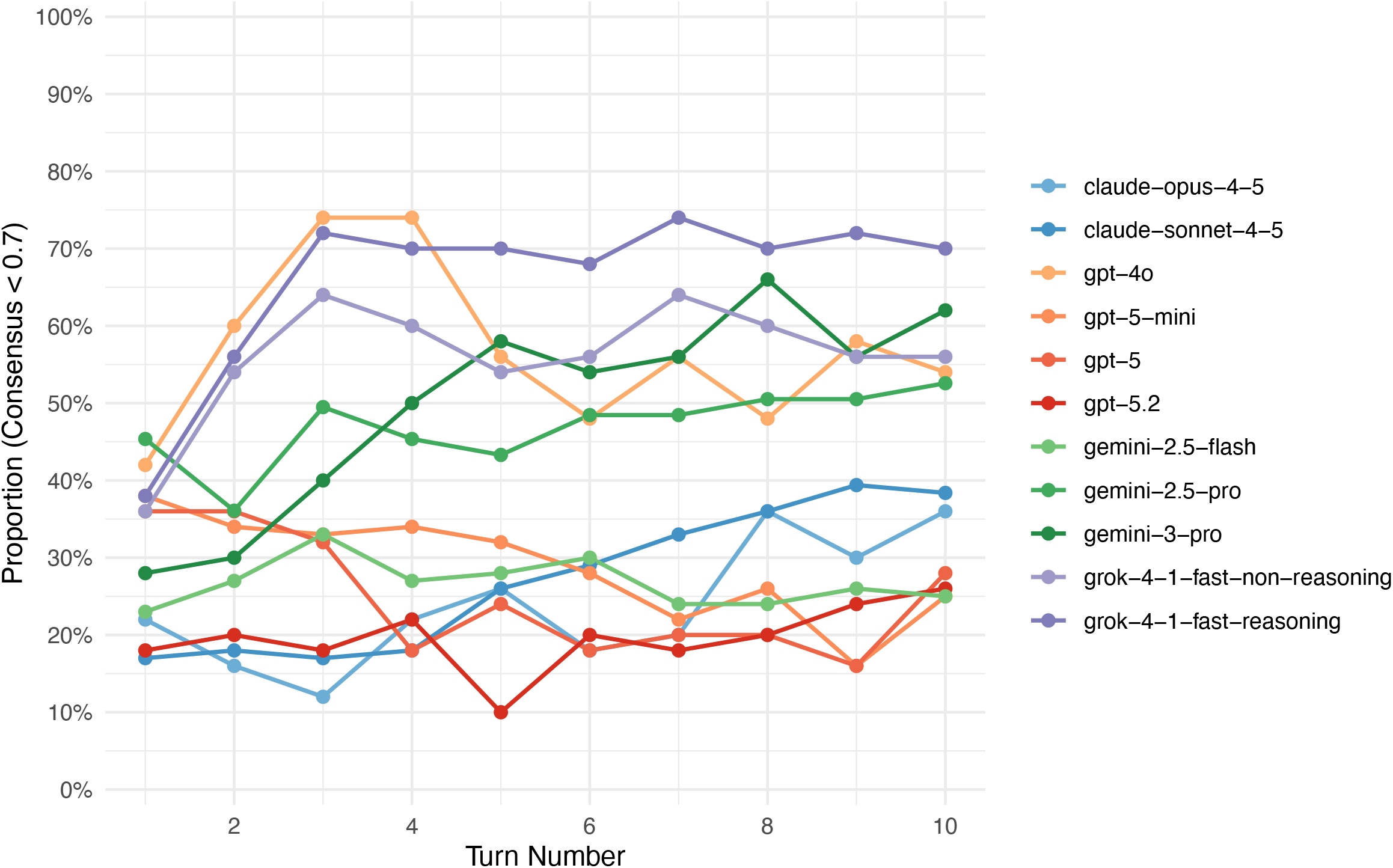
Proportion of Turns with Helpfulness Score < 0.7 (Pooled over all Scenarios)

## 4. Discussion

We developed and applied a configurable simulation harness to observe how frontier general-purpose chatbots behave across multi-turn mental-health conversations. By pairing persona-based simulated users with crisis and help-seeking scenarios, and by pre-specifying simple safety rubrics applied by an ensemble of judges, we were able to quantify how often models responded appropriately to acute risk, including both failing to respond harmfully, and responding helpfully, over repeated exchanges. We found that models that are widely deployed for general use failed safety criteria in a non-trivial fraction of high-risk conversations, and that these models exhibited substantial variation in performance by these metrics. These findings underscore that high average performance on generic benchmarks does not guarantee safe behavior in health-specific contexts.

Our results align with and extend recent audits of LLM responses to suicide-related queries and other high-risk mental-health content^12–14^. Multi-model evaluations examining a broader set of risk scenarios similarly report gaps in risk recognition, variability in escalation to crisis resources, and occasional provision of potentially harmful information^15,16,27^. Within this developing literature, the present work adds capability to conduct simulated randomized trials or large-scale cohort studies, probing model behavior across multi-turn encounters rather than single prompts or brief scripts. Results from multi-turn analyses suggest that risk does not diminish after initial responses, and for some models may increase with such multi-turn exchanges. Our test harness can be viewed as one way to implement and integrate emerging frameworks for evaluating these sorts of interactions.

An important strength of the approach we describe is the capacity to extend evaluations beyond simple pass–fail metrics for individual prompts. As conversations are structured as sequences of turns within simulated cohorts, it is possible to examine (for example) time-to-first-escalation using survival-analytic methods, to quantify how quickly different models move from risk recognition to explicit crisis referrals. The same infrastructure can be used to track drift over time by repeating identical cohorts at regular intervals as underlying models or guardrails change, to run counterfactual fairness tests by holding content constant while varying demographic attributes, and to evaluate localization by assessing whether models route users to country-appropriate crisis resources across languages. The platform can also randomize guardrail strategies—such as safety prompts or hotline-directory tools—enabling guardrail randomized controlled trials that estimate effect sizes for proposed safety interventions. Finally, its capacity for near-real-time iteration renders it appropriate for implementation as ongoing monitoring for chatbots, rather than as a single point estimate of safety.^19^ Ultimately, we envision continuous background simulation to detect low-frequency but high acuity harms, paired with real-time monitoring of actual conversations using similar metrics. In aggregate, this strategy approximates the postmarketing surveillance required of medications and medical devices, which may reach the market without a full understanding of their real-world safety.

## Limitations

This work should be considered with multiple limitations in mind. First, while it allows a broad range of simulated users, it cannot be considered to be a replacement for human testing. As with other forms of AI in medicine, maintaining humans in the loop will be critical in ensuring safety as chatbots are developed and deployed more generally, a need recognized even by the developers of the first therapy-focused chatbot in 1966^28^. A critical element of the implementation we describe is the capacity to flag and annotate exchanges where risk is identified. While concordance between an expert human annotator and model judgment is already substantial, we anticipate that, with further iteration, it should be possible to achieve even greater agreement. Second, while we incorporate standard guidance regarding specific clinical scenarios, what represents best practice in mental health is likely to continue to develop along with the underlying technologies. As such, continued efforts to refine and standardize guidance will be critical in maximizing the safety and utility of chatbots in mental health and health more generally.

## Conclusion

The architecture we describe allows efficient assessment of safety, efficacy, and engagement in a health context. Beyond informing model selection and development, this kind of test harness could support a learning-health-system approach to health chatbots. These strategies could help to ensure that both frontier model chatbots, and more focused health coaching and education applications, deliver maximal benefit while minimizing the risk of harm among vulnerable individuals.

## Data Availability

All summary data produced in the present study are available upon reasonable request to the authors

## Acknowledgement and Disclosures

The authors report no additional funding to support this work. Dr. Perlis reports compensation for service on scientific advisory boards for Atella.ai, Circular Genomics, Genomind, and Alkermes. The other authors are employees of Atella.ai and report no other competing interests.

